# Effect modification by sex of genetic associations of vitamin C related metabolites in the Canadian Longitudinal Study on Aging

**DOI:** 10.1101/2024.04.03.24305226

**Authors:** Rebecca Lelievre, Mohan Rakesh, Pirro G Hysi, Julian Little, Ellen E Freeman, Marie-Hélène Roy-Gagnon

## Abstract

Vitamin C is an essential dietary factor. There have been observed sex differences in serum vitamin C concentrations but the reason for this is not fully known. To understand how environmental factors like vitamin C intake interact with molecular processes, levels of metabolites can be used. Two metabolites associated with vitamin C are O-methylascorbate and ascorbic acid 2 sulfate. Past research has found there are genetic factors that influence these metabolite levels. Here, we aimed to investigate if there is effect modification by sex of these gene-metabolite associations and characterize the biological function of these interactions. We included individuals of European descent from the Canadian Longitudinal Study on Aging with available genetic and metabolic data (n= 9004). We conducted a genome wide association study with and without a sex interaction using mixed linear models. We also investigated the biological function of the important gene-sex interactions found for each metabolite. Two genome-wide statistically significant (p-value < 5×10^-8^) interaction effects and several suggestive (p-value < 10^-5^) interaction effects were found. The suggestive interaction associations were mapped to several genes including *HSD11B2*, which is associated with sex hormones and *AGRP* that helps initiate hunger drive. By understanding the genetic factors that impact metabolites associated with vitamin C, we better understand its function in disease risk. In addition, highlighting genetic markers and genes whose effects are modified by sex can help to understand the mechanisms behind sex differences in vitamin C levels and guide further research.

## Introduction

Vitamin C consumption and its effects on aging have been extensively studied^1,2^. There has been evidence to suggest that older adults do not consume a high enough concentration, and this may lead to increased risk of frailty and other conditions due to the immune effects of the vitamin^2,3^. Past research has also found differences between serum vitamin C concentrations in males and females, with males more likely to have lower concentrations^4,5^. Several theories have been proposed to account for this difference, including external factors such as body size and lifestyle factors and internal or molecular factors such as sex hormones^6^. Some studies which have noted the influence of sex hormones on vitamin C concentrations have relied on cellular and animal models^6–8^. One recent cross-sectional study in infertile men found there was an inverse association between serum ascorbic acid and luteinizing hormone levels, however, the relationship between vitamin C and sex hormones in humans needs to be further explored^9^.

The exposome is the totality of all environmental exposures in someone’s lifetime and how they influence disease^10^. One way to quantify a portion of the exposome is through the levels of metabolites. A metabolite refers to a substance that is produced or consumed during metabolism. Metabolites are intermediates in a metabolic pathway and can be substrates or products. Quantifying metabolites can bring insight into mechanisms by which the environment affects biological/metabolic processes^11^. For example, a study by Hysi et al., 2019 used this strategy to look at the metabolic associations of intraocular pressure (IOP) which is an endophenotype for glaucoma^12^.

Genetic factors can influence the levels of metabolites^13^. Chen et al., 2023^14^ recently conducted a large-scale genome-wide association study (GWAS) to identify genetic factors associated with all metabolites measured in participants of the Canadian Longitudinal Study on Aging (CLSA)^15,16^. Among the metabolites investigated were O-methylascorbate and ascorbic acid 2 sulfate, both of which are significantly associated with these two metabolites^14^. However, it is not known whether these genetic associations are affected by sex, which may explain some of the sex differences in vitamin C concentrations seen between males and females.

In this paper we aimed to investigate how sex affects the genetic association of variants across the genome with O-methylascorbate and ascorbic acid 2 sulfate in the CLSA data. In addition, using the comprehensive functional annotation platform FUMA^19^, we aimed to uncover the functional consequences of these genetic associations. This would help to investigate potential molecular mechanisms for sex-differences in vitamin C concentrations such as through an association with sex hormones. Understanding how genetic factors affect vitamin C-related metabolites can aid in understanding the influence of vitamin C on metabolic processes, and in turn how vitamin C influences disease phenotypes.

## Methods

### Study Population

We used baseline data from the Comprehensive Cohort of the Canadian Longitudinal Study on Aging (CLSA)^15^. The CLSA recruited participants between 2010 and 2015 who were between the ages of 45-85 years to investigate social, environmental, and other factors that affect aging and disease. The Comprehensive Cohort included 30 097 participants with baseline data collected between 2012 and 2015 via in-home interviews and in-person physical examinations and biospecimen sample collections at CLSA data collection sites located in Victoria, Vancouver, Surrey, Calgary, Winnipeg, Hamilton, Ottawa, Montreal, Sherbrooke, Halifax, and St. John’s, Canada. Inclusion criteria required participants to be community dwelling, be cognitively unimpaired, and to speak English or French. Not included were full-settlement, those living in a long-term care institution, and non-residents or citizens of Canada.

Of the Comprehensive Cohort, 9992 participants had metabolite levels quantified and 26 662 individuals were genotyped^16,20^. In this study we focused on ∼9000 CLSA participants of European ancestry with genetic and metabolic information and without any missing covariate information. Written informed consent was obtained for all participants, and research ethics board approval was obtained for all CLSA affiliated sites. The analysis presented here was approved by the University of Ottawa research ethics board.

### Genomic Data Quality Control

Blood samples were collected from consenting participants of the CLSA Comprehensive Cohort, and samples were moved to −80°C storage before shipment to the genomics facility where they were stored at −20°C. The Affymetrix Axiom array was used to perform genome-wide genotyping, resulting in 794,409 variants from 26,622 participants^20^. We followed the genetic ancestry procedures performed by the CLSA to identify participants of European descent^20^. The CLSA genomic data release included genotype data imputed using the TOPMed reference panel, resulting in ∼308 million variants imputed^21^. Prior to GWAS, we filtered out variants from the imputed data that had a minor allele frequency (MAF) <0.01, an imputation quality score <0.3 and missingness >0.1. After these filters, 8,836,359 variants remained for analysis. Both single nucleotide polymorphisms (SNPs) and insertions/deletions (INDELs) were included.

### Metabolite Processing

Metabolite levels were quantified using mass spectrometry and then identified using the Metabolon Discovery HD4TM LC-MS platform^16^. Metabolite values underwent quality control measures and 1314 metabolites were included in the final dataset. We were interested in 2 metabolites: O-methylascorbate and ascorbic acid 2 sulfate. In this GWAS we used the CLSA data with batch normalized values and where missing values were imputed with the lowest value recorded. Other researchers have reported using this imputation approach for missing values, and we assumed missing values were due to the limit of detection of the Metabolon platform^22^. For O-methyl ascorbate, there were no missing values, and for ascorbic acid 2 sulfate, there were 113 missing values. Prior to analysis, metabolites levels were log-transformed and extreme outliers (more than 3 SD away) removed followed by normalization to a mean of 0 and an SD of 1 which was done in prior studies^14^.

### Overall Genome-Wide Association Study

We first performed a GWAS on the complete dataset using mixed linear models as implemented in the GCTA/fastGWA program^23^. We used a sparse genetic relatedness matrix as a covariance structure to control population stratification and relatedness. We also adjusted models for age, sex, batch number, the first ten genetic principal components, province, and hours since the last meal or drink. After removing individuals with missing covariate values and outlier metabolites, there were 8916 participants for the O-methylascorbate GWAS and 8835 participants for the ascorbic acid 2 sulfate GWAS.

Manhattan plots and qqplots were made using the qqman^24^ package in R to visualize for any statistically significant variants. Results were filtered for p-values at the suggestive level (1 x 10^-5^) and the statistically significant level (5 x 10^-8^).

Independent (i.e. in linkage equilibrium) genetic variants were obtained using the GCTA/COJO program^25^ which implements a stepwise selection procedure to identify variants within significantly associated genomic regions that remain independently associated with the trait after conditioning on most statistically significant variants. The program also incorporates linkage disequilibrium (LD) structure information from an input population, which was set as the same individuals as those used in the GWAS analysis. Significant variant threshold was based on the genome-wide significance level of 5 x 10^-8^.

We investigated functional significance using the platform FUMA^19^ for lead variants identified by COJO analysis. To identify associated genes, positional mapping, quantitative expression quantitative trait loci (eQTL) mapping, and chromatin interaction mapping were used. FUMA identifies all variants in LD (based on 1000 Genomes LD structure) with lead variants to use for mapping. Variants were filtered prior to mapping to only those with a Combined Annotation Dependent Depletion (CADD)^26^ score above a threshold established as associated with variants with deleterious effects^19,27^ that based on research classifying the pathogenicity of genetic variants was set at the suggested level of >12.37^27^. For positional mapping, we additionally only used exonic or splicing variants. For eQTL mapping, a false discovery rate (FDR) threshold of <0.05 was adopted. For chromatin interaction mapping the threshold was an FDR of <1 x 10^-06^, in line with the default FUMA parameters^19^. All tissues were used for mapping.

### Gene-Sex Interaction GWAS

We performed a GWAS to test whether genetic effects were modified by chromosomal sex. To achieve this, we used the GCTA program fastGWA-GE and ran a mixed linear model adjusted for age, batch number, province, hours since last meal and the first 10 principal components^28^. We used the same sparse GRM and filtered for MAF 0.01 using the GCTA program, leading to 8,580,042 variants.

### Sex-Stratified GWAS

We also performed a sex stratified GWAS for each metabolite, adjusting for the same covariates as the overall GWAS. For O-methylascorbate, the number of participants was 4580 and 4329 for males and females, respectively. For ascorbic acid 2 sulfate, the number of participants was 4310 and 4518 for males and females, respectively.

### Finding Significant Signals of Interaction & Functional Consequences of Interaction Signals

We followed up all suggestive signals (p-value of interaction test < 1 x 10^-05^). We used the FUMA application to select lead variants and define loci of interest. The same parameters were used as before, such as filtering by CADD score; however, lead variants were not provided to FUMA externally. In this case, the application selects lead variants and independent significant variants based on LD (r^2^) information^19^. In brief, all variants with p-values below the suggestive threshold and independent from each other (r^2^ <0.6) were identified. Variants that were in r^2^ ≥ 0.6 with these variants and had a p-value less than 0.05 were equally considered for gene mapping. Lead variants were chosen from the identified suggestive variants if they had r^2^ <0.1 with other shortlisted variants.

In a small number of cases (8), variants that were suggestive were not recognized in the FUMA application, and required an alternative investigation. These variant regions were visualized using LD Link^29^ and lead variants and any additional annotated variants were selected based on similar criteria to FUMA. The CADD score of resulting variants was determined to filter variants that could be used for mapping. None of the identified variants were above the CADD threshold for deleteriousness and therefore were not mapped to genes. To annotate the functions of the lead variants and variants that met FUMA criteria, but which were not recognized in the platform, the Variant Effect Predictor (VEP) platform from Ensembl was used^30^.

To illustrate the direction of effect for each lead variant stratified by sex, we plotted interaction graphs using the results from the GWAS stratified by sex. We plotted the effect size of each lead variant by sex with a 95% confidence interval using R/ggplot. To report the results of this study, we were informed by Strengthening the Reporting of Genetic Associations (STREGA) guidelines^31^.

## Results

### Overall GWAS Results

Using the GCTA-fastGWA program we found 592 statistically significantly associated variants (p value < 5 x 10^-8^) with O-methylascorbate. For ascorbic acid 2 sulfate, we found 56 statistically significantly associated variants (p value <5 x 10^-8^). The Manhattan plot of these results is displayed in Figure S1 and Figure S2. The full list of suggestive variants and p-values for O-methylascorbate and ascorbic acid 2 sulfate are found in Table S1 and Table S2 respectively.

Using the GCTA-COJO software, we identified 3 independent lead variants for O-methylascorbate, all of which were on chromosome 22. For ascorbic acid 2 sulfate, we identified 2 lead variants on chromosome 16 and 1 lead variant at chromosome 10. The variant positions, effect allele frequencies, GWAS p-value and COJO-adjusted p-value are summarized in Table 1.

**Table 1.**
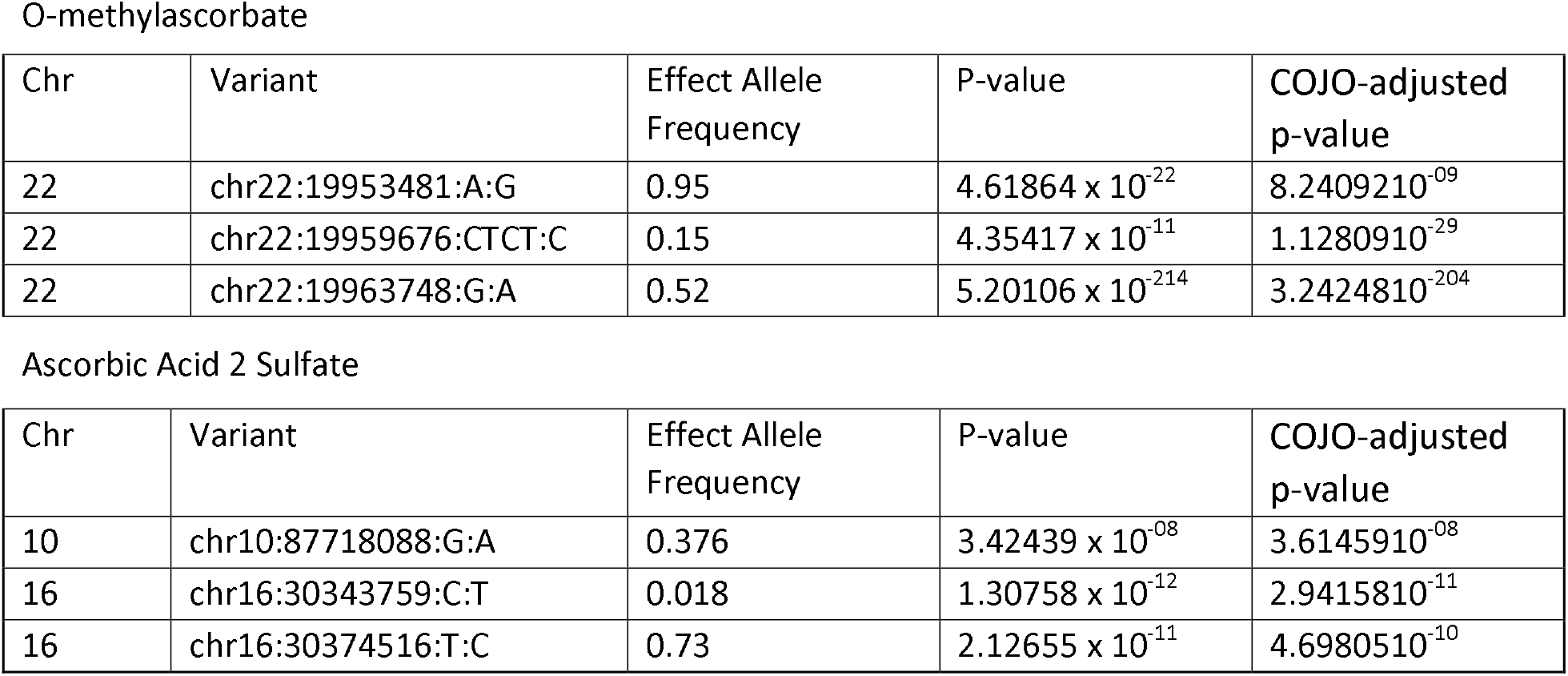
Lead variants from overall GWAS analysis selected using GCTA-COJO. Details of chromosome, position, effect allele frequency, p-value from GWAS, and COJO-adjusted p-value. Variants were selected using the GCTA-COJO program.

| Chr | Variant | Effect Allele Frequency | P-value | COJO-adjusted p-value |
| --- | --- | --- | --- | --- |
| 22 | chr22:19953481:A:G | 0.95 | $4.61864 \times 10^{-22}$ | $8.2409210^{-09}$ |
| 22 | chr22:19959676:CTCT:C | 0.15 | $4.35417 \times 10^{-11}$ | $1.1280910^{-29}$ |
| 22 | chr22:19963748:G:A | 0.52 | $5.20106 \times 10^{-214}$ | $3.2424810^{-204}$ |

Ascorbic Acid 2 Sulfate
| Chr | Variant | Effect Allele Frequency | P-value | COJO-adjusted p-value |
| --- | --- | --- | --- | --- |
| 10 | chr10:87718088:G:A | 0.376 | $3.42439 \times 10^{-08}$ | $3.6145910^{-08}$ |
| 16 | chr16:30343759:C:T | 0.018 | $1.30758 \times 10^{-12}$ | $2.9415810^{-11}$ |
| 16 | chr16:30374516:T:C | 0.73 | $2.12655 \times 10^{-11}$ | $4.6980510^{-10}$ |

### Functional Consequences of Genes

Using the FUMA platform, we determined the functional consequences of the lead variants found in the COJO analysis and variants in LD with those variants which met FUMA criteria. Variants associated with O-methylascorbate were mapped to 13 genes, which included both known and novel gene associations (Table S5). We subsequently used the FUMA platform to identify the functional consequences of the mapped genes, such as their expression levels in different tissue types and whether the set of genes was enriched in any functional pathways. The mapped genes were not significantly differentially expressed in any tissue types (Table S6). The functional gene sets linked to some processes such as genes which are involved in a cancer cell-death evasion mechanism^32^. No gene sets could be directly related to vitamin C functions (Table S7).

Variants associated with ascorbic acid 2 sulfate were mapped to 71 genes (Table S8). Using the same process as before, we found that the mapped genes were not significantly differentially expressed in any tissue types (Table S9). The mapped genes were linked to gene sets for chromosomal and novel genes were mapped to ascorbic acid 2 sulfate as well.

### Gene-sex Interaction GWAS

We conducted a gene-by-sex GWAS and found that, for O-methylascorbate, there were no statistically significant interactions while there were 69 suggestive interaction variants. For ascorbic acid 2 sulfate, we found 2 statistically significant interaction variants and 83 suggestive interaction variants. The significant variants were rs1296721356 (*chr1:13354706*) and rs1301173408 (*chr1:13341668*). Manhattan plots visualizing the associations are shown in Figure 1. The full list of suggestive interaction variants and p-values for O-methylascorbate and ascorbic acid 2 sulfate are found in Table S3 and Table S4 respectively.

**Figure 1.**
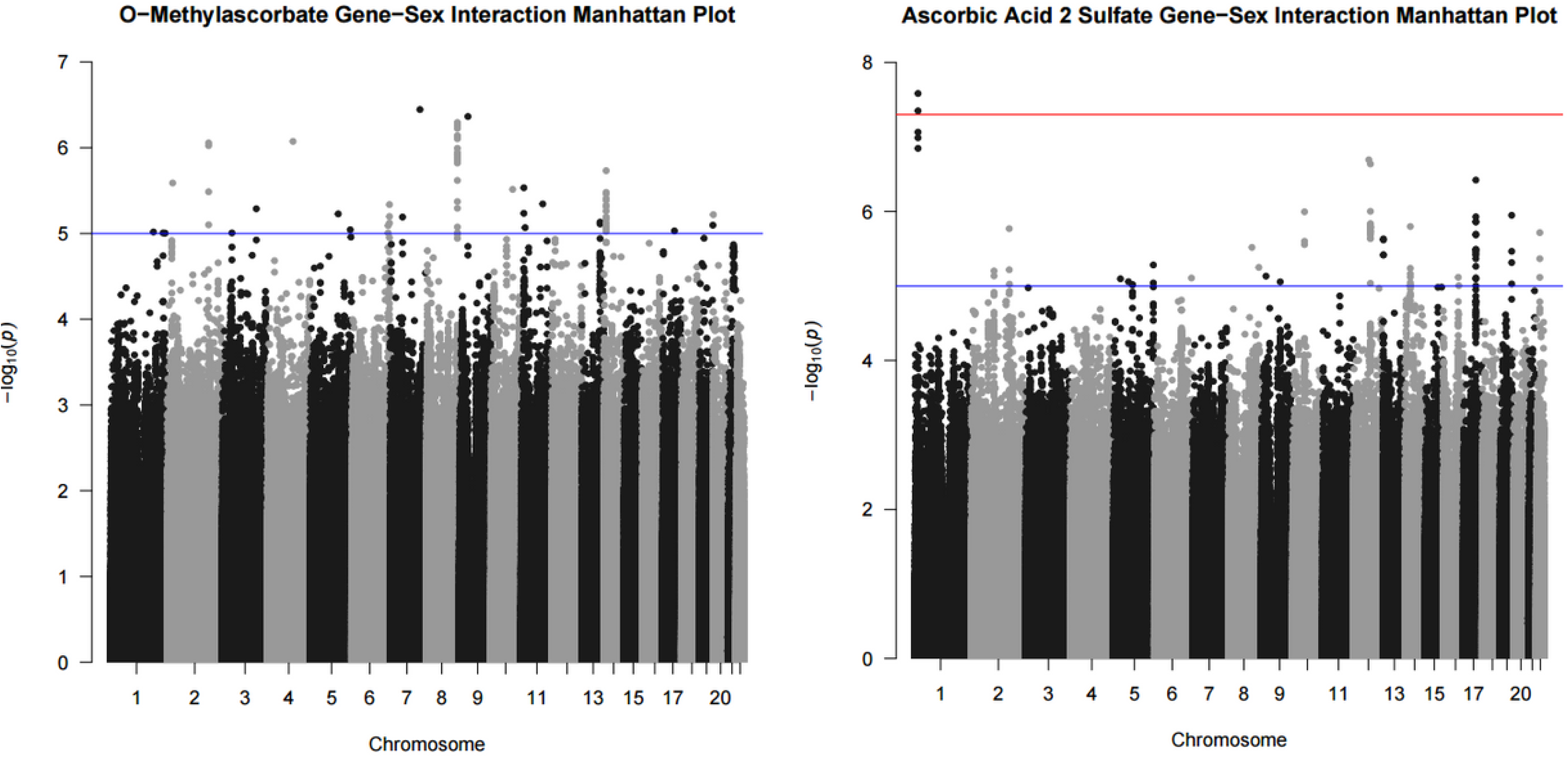
O-methylascorbate and ascorbic acid 2 sulfate interaction with sex results. Results from the GWAS analysis using the FastGWA-GE program from GCTA in the form of a Manhattan plot. GWAS conducted using a mixed linear model adjusted for age, batch number, province, 10 principal components, and hours since last meal or drink which incorporated a genetic relatedness matrix to account for population stratification. The models also included an interaction term for sex. Each point represents the p-value of the interaction between sex and the variant on the associated chromosome. The red line is the significant (5 x 10^-8^) threshold and the blue line is the suggestive (1 x 10^-5^) threshold.

We investigated all the signals at statistically significant and suggestive levels. Using the default parameters of FUMA, we identified 25 lead variants for O-methylascorbate and 23 lead variants for ascorbic acid 2 sulfate (Table S11, Table S12). After conducting separate sex stratified GWAS analyses for comparison, we saw that all interactions were qualitative in nature, meaning that the variants had opposite effects in males and females in the sex-stratified analyses (Figure 2).

**Figure 2.**
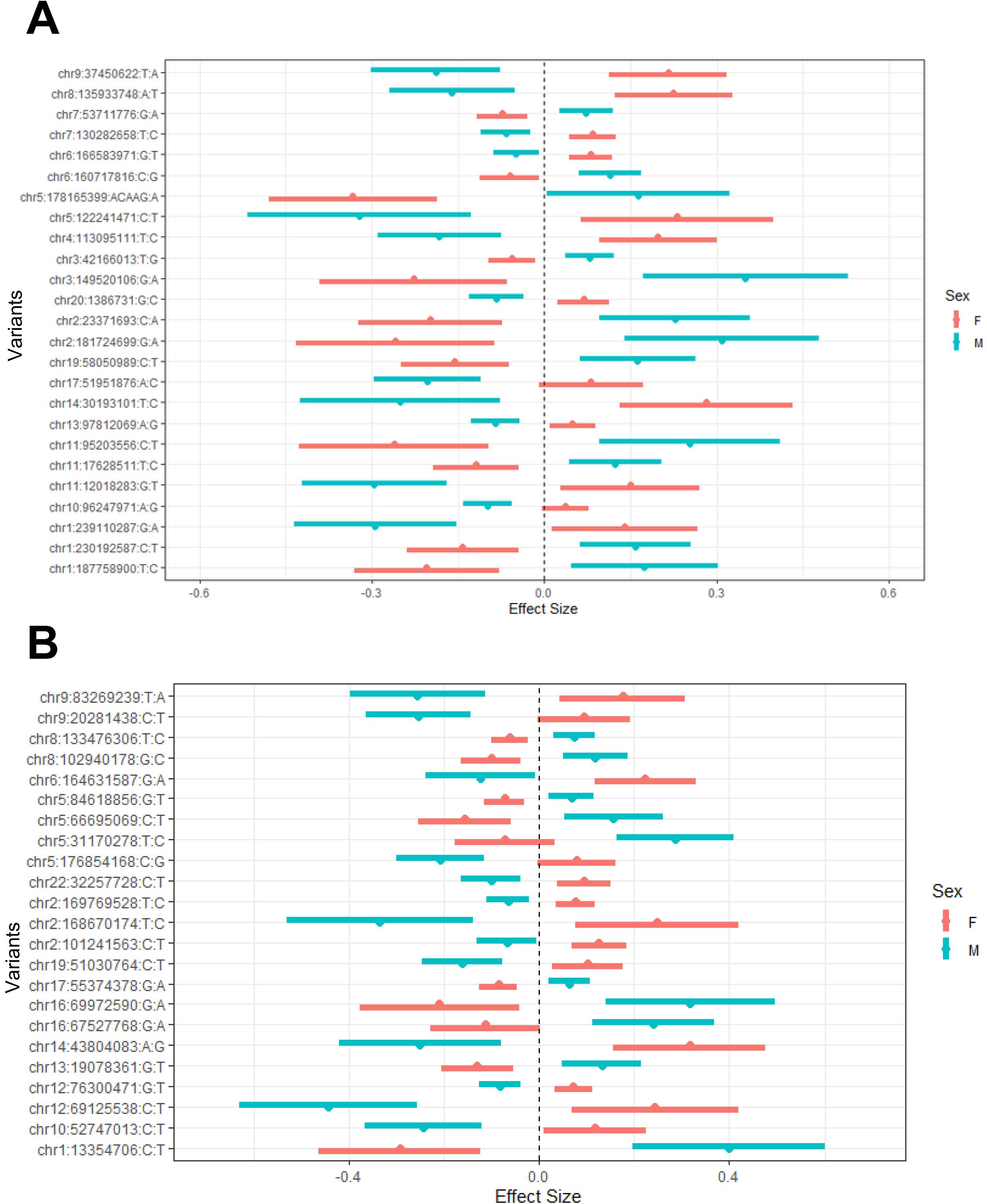
O-methylascorbate (A) and ascorbic acid 2 sulfate (B) effect sizes of lead interacting variant by sex. Effect sizes and 95% confidence intervals from the GWAS stratified by sex. GWAS conducted using a mixed linear model adjusted for age, batch number, province, 10 principal components, and hours since last meal or drink which incorporated a genetic relatedness matrix to account for population stratification. The list of variants are the lead variants for each suggestive locus found in the interaction GWAS analysis. Red represents female values and blue represents male values.

In Table 2 and Table 3, we summarized the lead variants for each suggestive interaction loci, nearest genes, annotated functions, and number of genes mapped at each suggestive genomic loci for O-methylascorbate and ascorbic acid 2 sulfate respectively. Overall, most of the genomic loci included variants in intronic or intergenic regions, which were not mapped to any genes. However, some regions had variants of likely functional importance which were mapped to more than one gene.

**Table 2.**
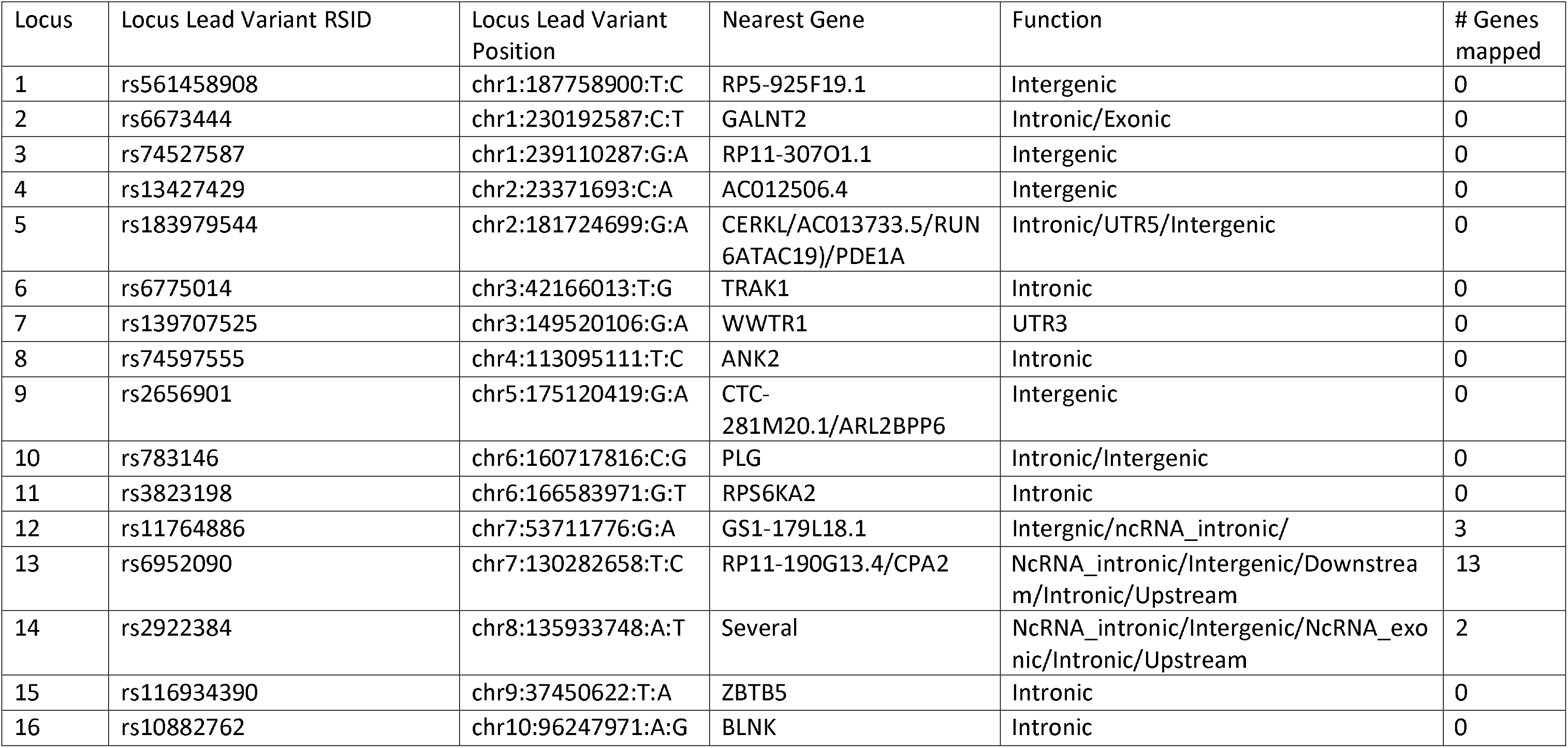
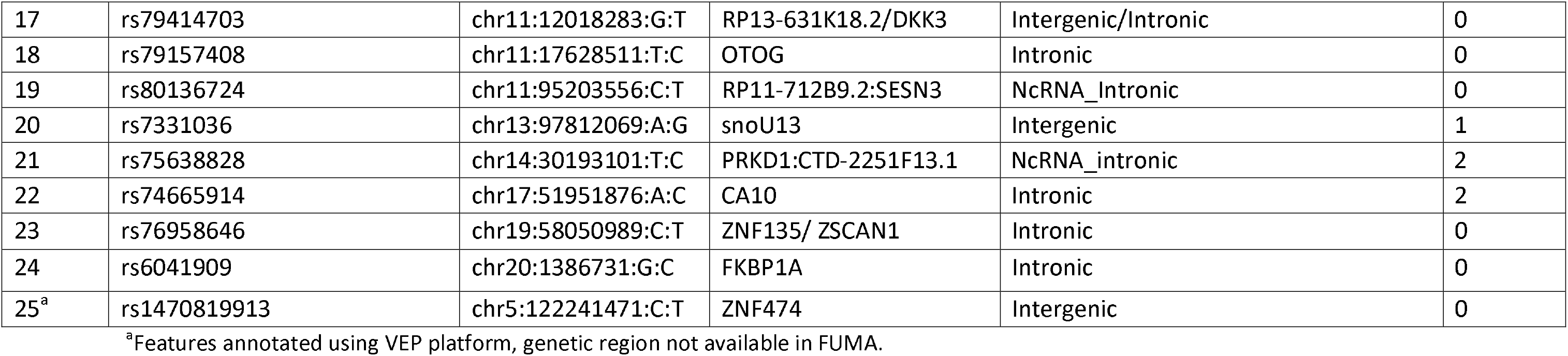
O-methylascorbate suggestive interaction loci functions. Lead variants for each suggestive interaction locus were selected based on FUMA criteria. For each locus, variants in LD of the lead variant were analyzed to note down the nearest gene and function. The number of genes mapped to each locus is based on FUMA criteria. Loci that did not register in FUMA, and whose functions and nearest gene information were retrieved using VEP are marked.

**Table 3.**
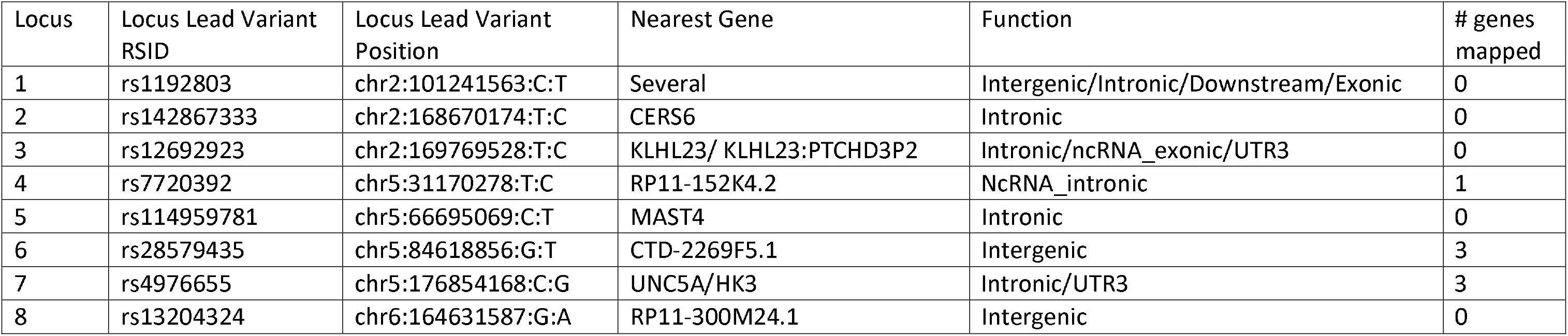
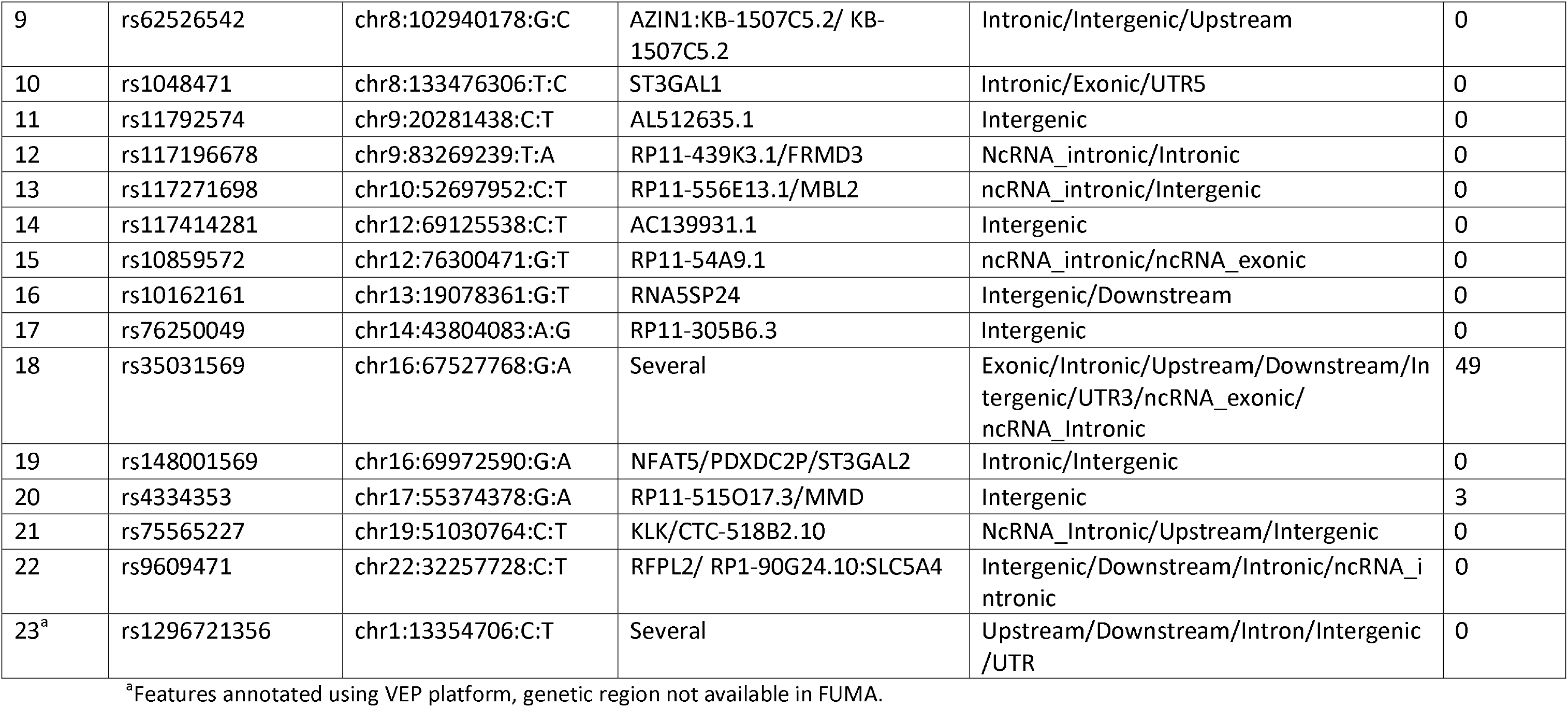
Ascorbic acid 2 sulfate suggestive interaction loci functions. Lead variants for each suggestive interaction locus were selected based on FUMA criteria. For each locus, variants in LD of the lead variant here analyzed to note down the nearest gene and function. The number of genes mapped to each locus is based on FUMA criteria. Loci that did not register in FUMA, and whose functions and nearest gene information were retrieved using VEP are marked.

| Locus | Locus Lead Variant RSID | Locus Lead Variant Position | Nearest Gene | Function | # genes mapped |
| --- | --- | --- | --- | --- | --- |
| 1 | rs1192803 | chr2:101241563:C:T | Several | Intergenic/Intronic/Downstream/Exonic | 0 |
| 2 | rs142867333 | chr2:168670174:T:C | CERS6 | Intronic | 0 |
| 3 | rs12692923 | chr2:169769528:T:C | KLHL23/ KLHL23:PTCHD3P2 | Intronic/ncRNA_exonic/UTR3 | 0 |
| 4 | rs7720392 | chr5:31170278:T:C | RP11-152K4.2 | NcRNA_intronic | 1 |
| 5 | rs114959781 | chr5:66695069:C:T | MAST4 | Intronic | 0 |
| 6 | rs28579435 | chr5:84618856:G:T | CTD-2269F5.1 | Intergenic | 3 |
| 7 | rs4976655 | chr5:176854168:C:G | UNC5A/HK3 | Intronic/UTR3 | 3 |
| 8 | rs13204324 | chr6:164631587:G:A | RP11-300M24.1 | Intergenic | 0 |
| 9 | rs62526542 | chr8:102940178:G:C | AZIN1:KB-1507C5.2/ KB-1507C5.2 | Intronic/Intergenic/Upstream | 0 |
| 10 | rs1048471 | chr8:133476306:T:C | ST3GAL1 | Intronic/Exonic/UTR5 | 0 |
| 11 | rs11792574 | chr9:20281438:C:T | AL512635.1 | Intergenic | 0 |
| 12 | rs117196678 | chr9:83269239:T:A | RP11-439K3.1/FRMD3 | NcRNA_intronic/Intronic | 0 |
| 13 | rs117271698 | chr10:52697952:C:T | RP11-556E13.1/MBL2 | ncRNA_intronic/Intergenic | 0 |
| 14 | rs117414281 | chr12:69125538:C:T | AC139931.1 | Intergenic | 0 |
| 15 | rs10859572 | chr12:76300471:G:T | RP11-54A9.1 | ncRNA_intronic/ncRNA_exonic | 0 |
| 16 | rs10162161 | chr13:19078361:G:T | RNA5SP24 | Intergenic/Downstream | 0 |
| 17 | rs76250049 | chr14:43804083:A:G | RP11-305B6.3 | Intergenic | 0 |
| 18 | rs35031569 | chr16:67527768:G:A | Several | Exonic/Intronic/Upstream/Downstream/Intergenic/UTR3/ncRNA_exonic/ncRNA_intronic | 49 |
| 19 | rs148001569 | chr16:69972590:G:A | NFAT5/PDXDC2P/ST3GAL2 | Intronic/Intergenic | 0 |
| 20 | rs4334353 | chr17:55374378:G:A | RP11-515O17.3/MMD | Intergenic | 3 |
| 21 | rs75565227 | chr19:51030764:C:T | KLK/CTC-518B2.10 | NcRNA_intronic/Upstream/Intergenic | 0 |
| 22 | rs9609471 | chr22:32257728:C:T | RFPL2/ RP1-90G24.10:SLC5A4 | Intergenic/Downstream/Intronic/ncRNA_intronic | 0 |
| 23<sup>a</sup> | rs1296721356 | chr1:13354706:C:T | Several | Upstream/Downstream/Intron/Intergenic/UTR | 0 |

<sup>a</sup>Features annotated using VEP platform, genetic region not available in FUMA.

Variants *rs1470819913* and *rs1296721356* were not available in FUMA and were annotated separately. No genes were mapped to these regions, and the summary of their functions is included in Table 2 and Table 3.

### Functional Consequences of Interaction Signals

The variants that interacting with sex were significantly associated with O-methylascorbate levels, were mapped to 23 different genes (Table S13). Looking at tissue expression data, the set of genes is statistically significantly differentially expressed (corrected p-value <0.05) in testis (Table S14).

The variants that interacted with sex for ascorbic acid 2 sulfate were mapped to 59 genes (Table S15). Looking at tissue expression data, the set of genes is statistically significantly differentially expressed (correct p-value <0.05) in the stomach (Table S16). One of the mapped genes was *AGRP* which is a neuropeptide which controls feeding behavior as a stimulating hormone antagonist^33^. Another gene mapped was *HSD11B2*, which is an enzyme involved in cortisol metabolism, and which has been shown to be controlled by sex hormones^34^. The significant genomic loci identified on chromosome 1 was not mapped to any genes and we did not find evidence of a potential functional consequence of this region based on our criteria. For both O-methylascorbate and ascorbic acid 2 sulfate, no gene-sets related to vitamin C metabolism or sex hormones were identified (Table S17, Table S18).

## Discussion

In the present study, we investigated associations and the functional consequences of gene-metabolite associations for two specific metabolites related to vitamin C: O-methylascorbate and ascorbic acid 2 sulfate. In addition, our study is the first to investigate potential gene-sex interactions influencing these two metabolites by conducting a gene by sex GWAS. We also examined the functionality of these variants of interest and found some associations with hormone-related genes.

Past research has studied the genomic associations of these metabolite levels, and Chen et al. 2023, conducted their study using the CLSA metabolite and genomic data. We aimed to have a more related metabolites specifically. We report the same associations, with *rs144009214* for ascorbic acid 2 sulfate and *rs61484427* and *rs4680* for O-methylascorbate^14^. We also examined additional lead variants than was previously reported due to our less stringent significance threshold of 5 x 10^-8^ (not corrected for examining all measured metabolites). Chen et al. Also identified *COMT* as the closest protein coding genes to the O-methylascorbate variants, which we also identified using FUMA. Using this platform, we were able to find several novel genes that were mapped to our GWAS results and should be investigated more thoroughly. Past studies looking at ascorbic acid 2 sulfate gene-metabolite associations have mapped them to the genes *CD2DP2* and *MAPK3*, which were both found in our present study, in addition to others^14,35^.

The FUMA analysis from the initial GWAS identified several new genes that were associated with metabolite levels. However, this comprehensive evaluation did not provide any new evidence of mechanisms or pathways through which these gene associations influence metabolite levels. In addition, none of the novel genes associated could be connected to vitamin C metabolism.

Of the mapped genes, *COMT* is a known factor in vitamin C metabolism. O-methylascorbate is a product of O-methylation by the *COMT* gene^36^. Therefore, this work strengthens an existing connection and further emphasizes the potential importance of this gene in vitamin C metabolism.

Our analysis of a potential sex effect on the gene associations seen for each metabolite found 2 significant interactions and several suggestive interactions. The effect of sex is an important factor to consider in disease etiology, and in this case, the sex differences of vitamin C concentrations between males and females and not completely understood. One of the proposed theories for the sex differences seen between vitamin C concentrations in males and females is due to influence by sex hormones^6^. We connections to genes related to hormone signaling or sex hormones.

For both ascorbic acid 2 sulfate and O-methylascorbate, there were no gene-sets which are enriched and may be related to vitamin C metabolism or sex hormones. Interestingly, the set of genes mapped for ascorbic acid 2 sulfate was significantly differentially expressed in stomach tissues while the set of genes mapped for O-methylascorbate was differentially expressed in testis tissues.

The variants that interacted with sex for ascorbic acid 2 sulfate were mapped to several genes, one of which was *AGRP*, which produces a peptide agonist molecule important for initiating hunger cues. In general, the *AGRP* neurons signal for increased food intake^33^. In one animal study using a bird model, researchers found that this protein was differentially expressed between male and female chickens^37^. Since vitamin C intake is heavily influenced by dietary exposures, this association with a hunger-driving signal protein may be something of further consideration.

Other variants that interacted with sex for ascorbic acid 2 sulfate were mapped to *HSD11B2*. The role of11-β hydroxysteroid dehydrogenase type 2, the enzyme encoded by the *HSD11B2* gene, is to oxidize cortisol, a glucocorticoid, into its inactive version cortisone^38^. Several animal and in vivo studies have shown that the activity of this enzyme is regulated by various sex hormones^34,39,40^. In addition, cortisol may potentially affect vitamin C concentrations in the body; however, it is unclear whether this acts in a sex dependent manner^6^. Overall, this may point to an important mechanism for future research.

This study has several strengths, including the use of high quality genetic and metabolic datasets. In addition, the use of such a comprehensive tool to annotate variant functions allowed us to use several bioinformatics tools to identify novel associations and areas for future research.

This study also had some limitations to consider. One limitation is the sample size, which may have hindered the ability to find more statistically significant associations, especially in the interaction analysis. However, these findings showed several associations at the suggestive level, which could be followed up by other researchers. Future studies should replicate these findings ideally with a larger sample size. Another limitation is that this study was only conducted using participants of European descent to avoid problems with population stratification. The CLSA had a very high percentage of participants of European descent so using populations from other ancestries would not provide enough power for an accurate analysis in those groups. Finally, because we decided to use the suggestive threshold for the interaction signals to follow up with for functional annotation and mapping, there is a possibility that some of these associations represent type I error. As this represents the first analysis of a potential sex interaction, future studies should evaluate these regions with larger sample sizes to determine if they are true associations.

In conclusion, our study found potential evidence for an effect modification by sex of genetic associations with two vitamin C related metabolites. In addition, a comprehensive analysis of the functions of genomic regions showing suggestive evidence of gene-sex interactions led to some insight into potential mechanisms for these differences. Future studies are needed to expand on this analysis and further understand the different mechanisms which influence vitamin C concentrations in the body. Mechanisms influencing vitamin C concentrations have several implications for different disease phenotypes^41,42^ and are an especially important consideration for older adults.

## Data Availability

Data are available from the Canadian Longitudinal Study on Aging (www.clsa-elcv.ca) for researchers who meet the criteria for access to de-identified CLSA data. Code is made available on GitHub (https://github.com/Roy-Gagnon-lab). Summary statistics for the interaction GWAS will be made available on GWAS Catalog (Data are available from the Canadian Longitudinal Study on Aging (www.clsa-elcv.ca) for researchers who meet the criteria for access to de-identified CLSA data. Code is made available on GitHub. Summary statistics for the interaction GWAS will be made available on GWAS Catalog.

https://www.ebi.ac.uk/gwas/

https://github.com/Roy-Gagnon-lab

## Supplemental Information

The supplemental information includes 2 figures and 18 tables.

## Declarations of Interests

The authors declare no competing interests.

## Acknowledgements

Funding for the Canadian Longitudinal Study on Aging (CLSA) is provided by the Government of Canada through the Canadian Institutes of Health Research (CIHR) under grant reference: LSA 94473 and the Canada Foundation for Innovation, as well as the following provinces, Newfoundland, Nova Scotia, Quebec, Ontario, Manitoba, Alberta, and British Columbia. This research was funded by CIHR operating grant PJT-180615 to Drs Roy-Gagnon and Freeman. The funders had no role in the design, analysis, or the interpretation of results.

This research was made possible using the data/biospecimens collected by the Canadian Longitudinal Study on Aging (CLSA). This research has been conducted using the CLSA Comprehensive Baseline Dataset version 7.0, the CLSA Metabolomics dataset version 1.0 and the CLSA Genomic dataset version 3.0 under Application Number 180911. The CLSA is led by Parminder Raina, Christina Wolfson, and Susan Kirkland. The opinions expressed in this article are the authors’ own and do not reflect the views of the Canadian Longitudinal Study on Aging.

The authors thank Dr. Julie St-Pierre for helpful comments regarding the metabolomics data.

## Data and code availability

Data are available from the Canadian Longitudinal Study on Aging (www.clsa-elcv.ca) for researchers who meet the criteria for access to de-identified CLSA data. Code is made available on GitHub

(https://github.com/Roy-Gagnon-lab). Summary statistics for the interaction GWAS will be made available on GWAS Catalog (https://www.ebi.ac.uk/gwas/).

## Notes

### Competing Interest Statement

The authors have declared no competing interest.

### Author Declarations

The analysis presented here was approved by the University of Ottawa research ethics board.

