## Supplemental Figure 1 for "Effect modification by sex of genetic associations of vitamin C related metabolites in the Canadian Longitudinal Study on Aging"

<u>Supplemental Information</u>

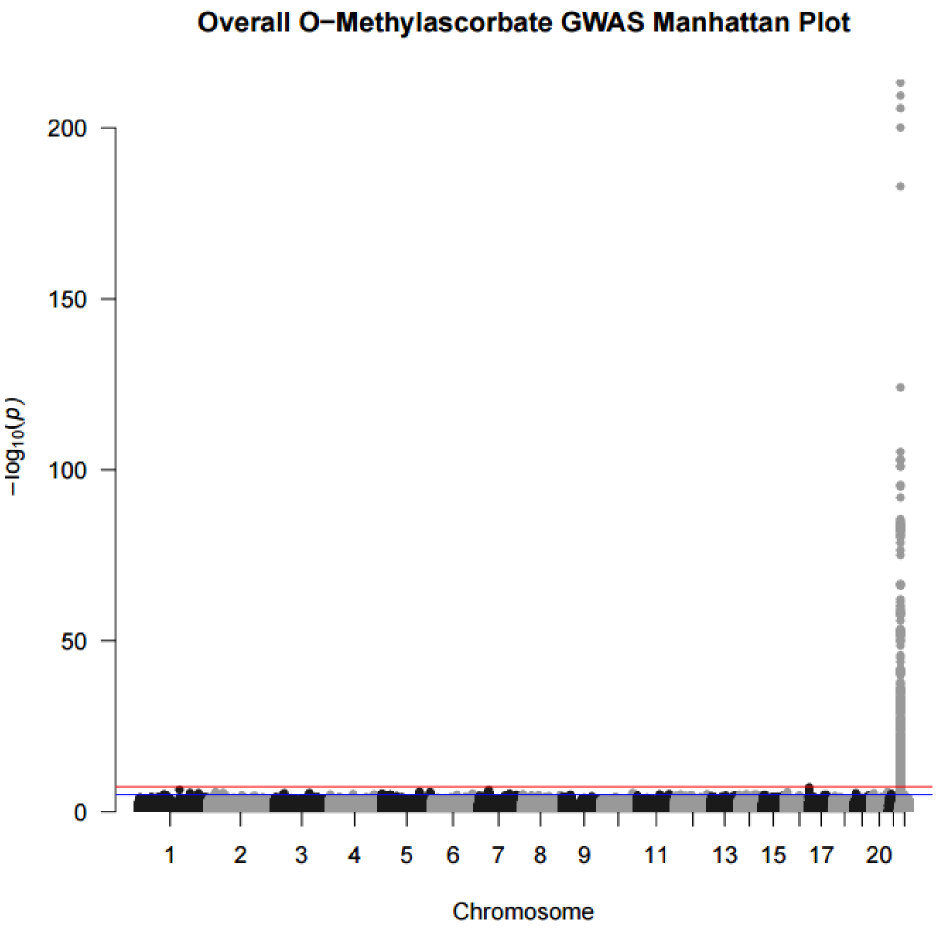


Figure S1. **O-methylascorbate** overall **results.** Results from the GWAS analysis using the FastGWA program from GCTA in the form of a Manhattan plot. GWAS conducted using a mixed linear model adjusted for age, sex, batch number, province, 10 principal components, and hours since last meal or drink which incorporated a genetic relatedness matrix to account for population stratification. Each point represents the p-value of a variant on the associated chromosome. The red line is the significant ($5 \times 10^{-8}$) threshold and the blue line is the suggestive ($1 \times 10^{-5}$) threshold.

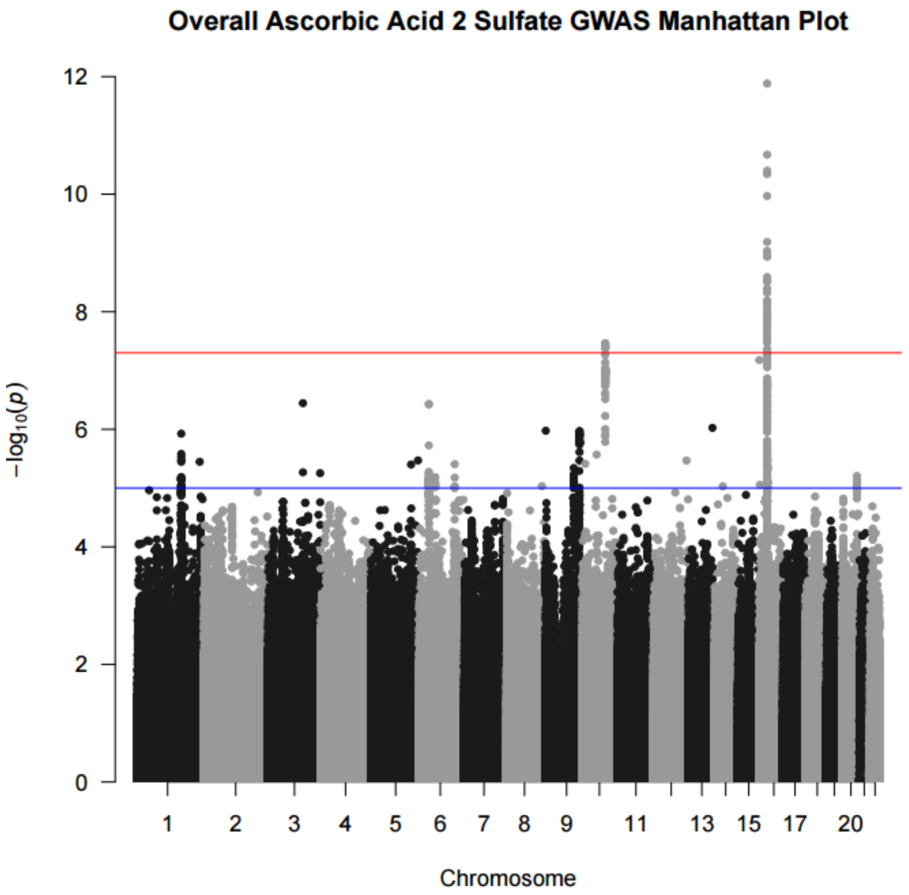


Figure S2. **Ascorbic acid 2 sulfate** overall **results.** Results from the GWAS analysis using the FastGWA program from GCTA in the form of a Manhattan plot. GWAS conducted using a mixed linear model adjusted for age, sex, batch number, province, 10 principal components, and hours since last meal or drink which incorporated a genetic relatedness matrix to account for population stratification. Each point represents the p-value of a variant on the associated chromosome. The red line is the significant ($5 \times 10^{-8}$) threshold and the blue line is the suggestive ($1 \times 10^{-5}$) threshold.
